# DIAGNOSTIC ACCURACY OF VISUAL INSPECTION WITH ACETIC ACID FOR CERVICAL CANCER SCREENING AMONG WOMEN LIVING WITH HUMAN IMMUNODEFICIENCY VIRUS IN OSOGBO, NIGERIA

**DOI:** 10.64898/2026.08.05.26359761

**Authors:** Sekinah B. Bola-Oyebamiji, Ibraheem Olayemi Awowole, Sunday Charles Adeyemo, Ayodeji Olaolu Oyeniran, Titilope A. Bamkefa, Bello Yussuf Olatunji, Daniel A. Adekanle, Eniola Dorcas Olabode

**Affiliations:** Obstetrics and Gynecology Department, Osun State University, Osogbo, Nigeria; Department of Obstetrics, Gynaecology and Perinatology, Obafemi Awolowo University, Ile-Ife, Nigeria; Health and Biomedical Sciences, Institut Superieur de Sante, Niamey, Niger Republic; Medicine Department, Osun State University, Osogbo, Nigeria; HPV consortium, College of medicine, University of Ibadan, Ibadan, Nigeria

**Keywords:** Cervical cancer screening, visual inspection with acetic acid, Human Papilloma Virus Women living with HIV, Nigeria

## Abstract

**Background:** Women living with Human Immunodeficiency Virus (WLHIV) are at high risk of cervical cancer. While Human Papilloma Virus Deoxyribonucleic acid (HPV DNA) testing is the standard of care, its cost limits widespread use in resource-limited settings, therefore visual inspection with acetic acid (VIA) remains the primary screening method. This study therefore evaluated the diagnostic accuracy of VIA compared to HPV DNA testing for cervical cancer screening among WLHIV in Nigeria.

**Methods:** This cross-sectional analytical study was conducted between November 2022 and November 2024 in Osogbo, Nigeria among 300 WLHIV on antiretroviral therapy aged 25-49 years underwent cervical cancer screening using VIA and HPV DNA testing (Ampfire HPV test kits, ATILA Biosystems® USA). All 309 participants underwent colposcopy and biopsy regardless of their screening results, with histological examination serving as the gold standard. Data were analyzed using Stata, with diagnostic performance measures calculated using histology as the gold standard.

**Results:** The mean age of participants was 42.6 ± 6.4 years. Thirty-six of 309 women (11.7%) were VIA-positive, while 91 (29.4%) were HPV-positive. Histology confirmed CIN2+ in 22 participants (7.1%). HPV testing demonstrated significantly higher sensitivity than VIA for detecting CIN2+ (91.2%, 95% CI: 76.3-98.1 vs. 44.1%, 95% CI: 26.7-62.6; p<0.001) and higher negative predictive value (98.9%, 95% CI: 96.3-99.8 vs. 92.0%, 95% CI: 88.0-95.0; p<0.001). However, VIA showed higher specificity (91.7%, 95% CI: 87.8-94.7 vs. 83.3%, 95% CI: 78.4-87.6; p=0.003). The agreement between the two tests was fair (kappa = 0.20, p<0.001). Both HPV positivity (adjusted OR: 42.1, 95% CI: 9.5-186.2; p<0.001) and VIA positivity (adjusted OR: 6.9, 95% CI: 2.6-18.4; p<0.001) were independently associated with CIN2+.

**Conclusion:** HPV DNA testing demonstrated superior sensitivity and negative predictive value compared to VIA for detecting CIN2+ among WLHIV, while VIA showed higher specificity.

## Introduction

Cervical cancer remains a disease of significant public health concern, being the fourth most common cancer worldwide, with approximately 604,000 new cases and 342,000 deaths in 2020 [1]. The World Health Organization (WHO) reported that over 80% of affected women reside in low- and middle-income countries (LMIC). In Nigeria, cervical cancer is the second most common cancer affecting women [2]. In 2020, 7,968 of the 12,075 women diagnosed with cervical cancer died from the disease [1].

Immunosuppression increases the risk for cervical cancer. Women living with HIV (WLHIV) have a six-fold increased susceptibility to cervical cancer compared to the general population [3]. This has been attributed to their increased susceptibility to HPV infection, rapid progression to cervical intraepithelial neoplasia (CIN), and an increased tendency towards recurrence [4]. HIV also aggravates the progression of CIN to cervical cancer due to persistence of HPV [3].

In Nigeria, 960,000 of over 1.7 million people living with HIV (56.5%) are women [5,6]. Considering that most people living with HIV are women, a nationwide structured screening program covered by the National Health Insurance Scheme ought to be available as part of the comprehensive care for WLHIV; however, this is not currently the case. Aid and support from high-income nations such as the United States provide routine care for WLHIV, but this approach is unsustainable [7].

In recognition of the importance of cervical cancer prevention, WHO developed a global strategy for the elimination of cervical cancer [8]. The WHO call for action involves vaccination of 90% of eligible young girls with the HPV vaccine, screening 70% of eligible women with a high-performance test such as HPV testing, and appropriate treatment in 90% of women with cervical pre-invasive disease and cancer by 2030 [8]. Adopting this strategy can potentially prevent 60 million deaths globally [9].

The standard of care for cervical cancer screening recommended by WHO is co-testing with cytology and HPV testing [8]. However, cost has limited the adoption of these screening methods in Nigeria and other LMICs. Instead, visual inspection with acetic acid or Lugol’s iodine (VIA/VILI) is commonly used because VIA is inexpensive, requires minimal training, and can be performed by nurses and other lower-level healthcare professionals [10], although it has been shown to have low sensitivity and persistent inter-observer variation [11, 12]. HPV testing is the test of choice in women living with HIV, as it is not associated with ambiguity in the interpretation of results [13–18].

Given that screening and early treatment improve cervical cancer outcomes, further comparative studies of VIA and HPV testing in WLHIV are warranted [19, 20]. VIA continues to find relevance in clinical practice in LMICs due to its low cost and minimal training requirements, despite HPV testing being the standard of care for WLHIV. However, a significant research gap exists regarding the comparative effectiveness of these tests in detecting precancerous lesions in resource-limited settings such as Nigeria, particularly as previous studies have been limited by verification bias. This study therefore aimed to compare the diagnostic accuracy of VIA with HPV DNA testing in diagnosing CIN using histology as the gold standard, with complete histological verification of all participants regardless of screening result, among WLHIV in Osogbo.

## Methods

### Study Design and Participants

This cross-sectional analytical study was conducted between November 2022 and November 2024 at the two largest public hospitals in Osogbo, Nigeria: Osun State University Teaching Hospital (UTH) and State Specialist Hospital, Asubiaro. It was carried out among women living with Human Immunodeficiency Virus (WLHIV). The inclusion criteria included WLHIV aged 25 to 49 years who had been on ART for at least 1 year, with no previous history of treatment for cervical intra-epithelial neoplasia. WLHIV with undiagnosed vaginal bleeding or vaginal discharge were excluded.

The two hospitals were purposively selected because they serve as major referral centres for WLHIV, provide comprehensive HIV care, and have the necessary infrastructure for colposcopy and biopsy, enabling complete histological verification of all participants. Participants were selected using simple random sampling. Ethical clearance was obtained from the Research Ethics Committee of UTH, Osogbo (UTH/REC/2022/05/17/603). Written informed consent was obtained from each participant before enrollment into the study.

### Sample Collection and Visual Inspection with Acetic Acid

Participants who consented to participate in the study underwent a pelvic examination using Cusco’s speculum to expose the cervix. A swab stick was inserted to obtain exfoliated cells and secretions for HPV screening. The transformation zone (TZ), delineated by the old and new squamo-columnar junction (SCJ), was identified using a light source. Five percent acetic acid was then applied to the cervix for 60 seconds. The presence of aceto-white lesions constituted a positive VIA test, while those without aceto-white changes were labelled VIA-negative. The VIA examinations were performed by trained research nurses who had undergone a standardized training program on VIA technique and interpretation specifically for this study. The training included theoretical sessions and practical demonstrations, with quality assurance measures to ensure uniformity across both study sites. Inter-observer reliability was assessed during training with a kappa statistic of 0.78, indicating substantial agreement.

### Human Papillomavirus Testing

Screening for HPV was performed using Ampfire HPV Screening 16/18/HR kits manufactured by ATILA Biosystems USA. The kit quantitatively detected HPV types 16, 18, 33, 35, 39, 45, 51, 52, 53, 56, 58, 59, 66, and 68 in cervical samples. The analysis was conducted at the Gynaecologic Oncology Research Laboratory of Obafemi Awolowo University Teaching Hospitals Complex, Ile-Ife.

### Colposcopy and Biopsy

All 309 participants underwent colposcopic evaluation by a trained colposcopist (a consultant gynaecologist with expertise in colposcopy), regardless of their VIA or HPV screening results. The colposcopist was blinded to the screening results to prevent interpretation bias. During the colposcopy, the cervix was painted with 5% acetic acid, and any area of aceto-whitening was biopsied. In women with no visible aceto-white lesions, a random biopsy was taken from the squamo-columnar junction to ensure histological verification. Histopathological examination was performed by a pathologist who was blinded to the screening test results. All women who had aceto-white lesions on colposcopy were referred for treatment.

Patients who tested positive for HPV 16/18 underwent thermo-coagulation. This treatment was performed after colposcopy and biopsy had been completed to avoid altering the histological findings. Lesions that occupied over 75% of the cervix or were noted to extend into the endocervical canal (exclusion criteria for thermo-coagulation) proceeded to loop electrosurgical excision procedure (LEEP) and further assessments.

### Statistical Analysis

Data were analyzed using Stata (StataCorp, College Station, TX, USA). Participants’ sociodemographic, reproductive, and clinical characteristics, awareness of cervical cancer, and screening history were summarized using frequencies and percentages. Overall concordance between VIA and HPV was examined using cross-tabulation, with results presented as frequencies and percentages. The strength of association between VIA and HPV results was assessed using the chi-square test, while agreement between the two tests was evaluated with Cohen’s Kappa statistics. The diagnostic performance of VIA and HPV testing in detecting CIN2+ lesions was summarized using sensitivity, specificity, positive predictive value (PPV), negative predictive value (NPV), and likelihood ratios, with histology as the gold standard. With complete histological verification of all 309 participants, unbiased estimates of diagnostic accuracy were obtained. Associations between participant characteristics and HPV infection were examined using the chi-square test. Logistic regression models were fitted to estimate crude and adjusted odds ratios (ORs) with 95% confidence intervals (CIs) for factors associated with cervical HPV infection. All statistical analyses were performed at a 5% level of significance (p-value < 0.05).

## Results

### Participant Characteristics

A total of 309 women participated in the study and all underwent complete histological verification. The mean age of participants was 42.6 ± 6.4 years, with the majority (67.0%) aged 41 years and above. Most participants were married (70.5%), had secondary education (39.6%), and were semi-skilled workers (36.7%). The mean parity was 3.8 ± 1.6, with the majority (56.7%) having 3-5 pregnancies. The mean age at menarche was 15.2 ± 2.1 years, and the mean age at first intercourse was 20.4 ± 3.8 years. Table 1 presents the full sociodemographic and reproductive characteristics of the participants.

**Table 1:** Participant Sociodemographic and Reproductive Characteristics.

| **Variable** | **Frequency (n=309)** | **Percentage** |
| --- | --- | --- |
| **Age at last birthday (years)** |  |  |
| 25-30 | 15 | 4.9 |
| 31-40 | 87 | 28.2 |
| 41 and above | 207 | 67.0 |
| **Marital Status** |  |  |
| Single | 14 | 4.6 |
| Married | 215 | 70.5 |
| Widowed | 55 | 18.0 |
| Separated | 21 | 6.9 |
| **Level of Education** |  |  |
| No formal education | 21 | 6.9 |
| Primary | 60 | 19.8 |
| Secondary | 120 | 39.6 |
| Tertiary | 102 | 33.7 |
| **Occupation** |  |  |
| Unskilled | 100 | 33.3 |
| Semi-skilled | 110 | 36.7 |
| Skilled | 63 | 21.0 |
| Professional | 27 | 9.0 |
| **Age at Menarche (years)** |  |  |
| ≤13 | 60 | 22.1 |
| 14-17 | 153 | 56.5 |
| 18 and above | 58 | 21.4 |
| **Age at First Intercourse (years)** |  |  |
| ≤15 | 10 | 3.7 |
| 16-20 | 124 | 46.1 |
| 21-25 | 101 | 37.6 |
| 26 and above | 34 | 12.6 |
| **Number of Pregnancies** |  |  |
| No previous pregnancy | 15 | 4.9 |
| 1-2 | 64 | 20.9 |
| 3-5 | 174 | 56.7 |
| 6 and above | 54 | 17.6 |
| **Number of Miscarriages/Abortions** |  |  |
| 0 | 172 | 57.1 |
| 1 | 77 | 25.6 |
| 2 and above | 52 | 17.3 |
| **Previous use of family planning method** |  |  |
| No | 168 | 55.1 |
| Yes | 137 | 44.9 |

### Screening Results

Of the 309 participants, 36 (11.7%) had positive VIA results, while 91 (29.4%) tested positive for HPV (Figure 1). Among the participants, 21 (6.8%) tested positive for both VIA and HPV, while 203 (65.7%) tested negative for both. However, 85 (27.5%) had discordant results between VIA and HPV testing. The overall agreement between VIA and HPV testing was fair, with a Cohen’s Kappa of 0.20 (p < 0.001) (Table 2). Among VIA-negative participants, 69 of 273 (25.3%) were HPV-positive.

**Figure 1:**
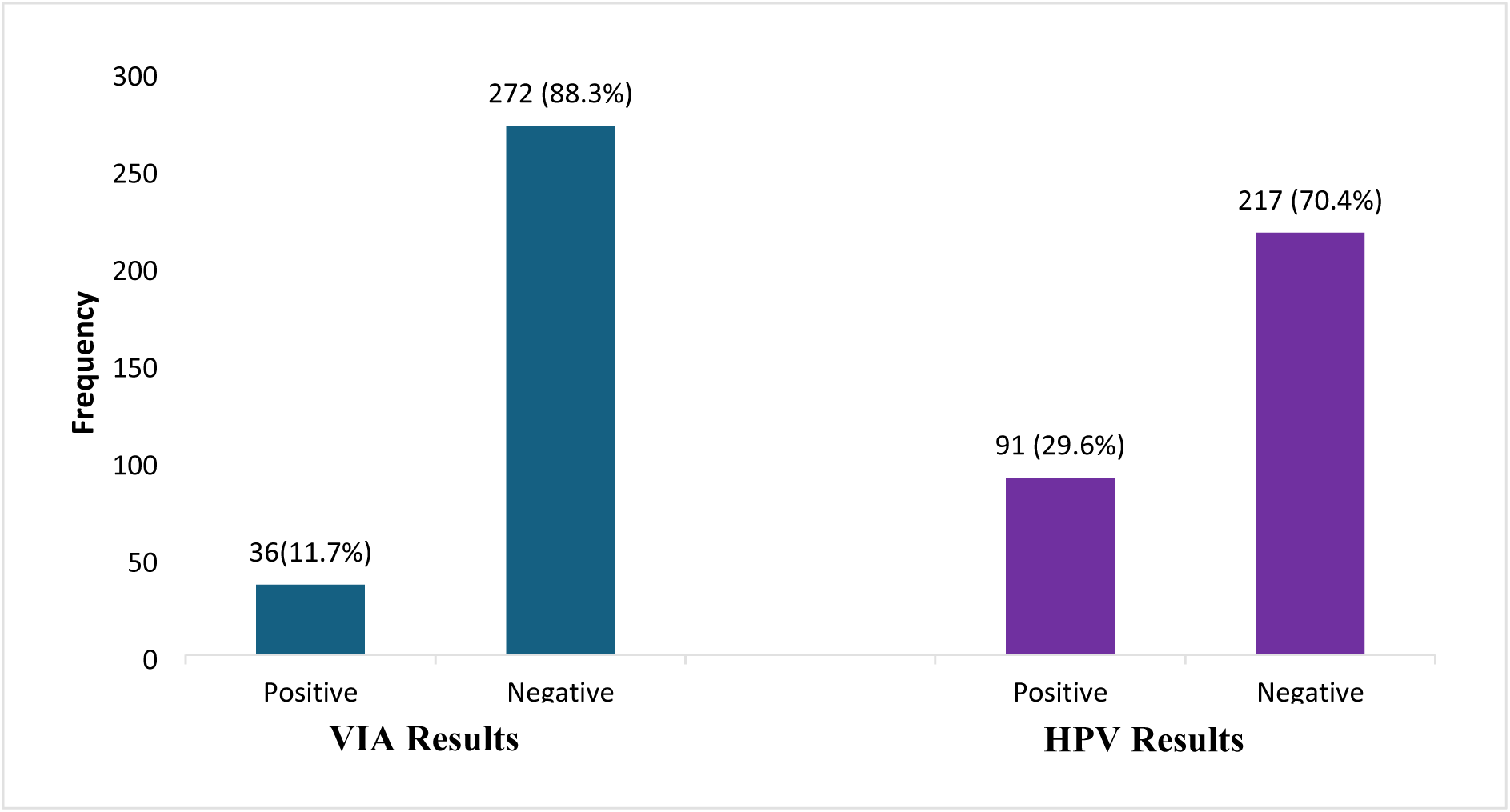
Distribution of VIA and HPV test results among study participants.

**Table 2:** Congruence between HPV Status and VIA Findings.

| Variable | HPV Positive n (%) | HPV Negative n (%) | Total | p-value |
| --- | --- | --- | --- | --- |
| **VIA Result** |  |  |  | **0.001** |
| Positive | 21 (6.8) | 15 (4.9) | 36 (11.7) |  |
| Negative | 69 (22.3) | 203 (65.7) | 272 (88.0) |  |
| Total | 90 (29.1) | 218 (70.6) | 308 (99.7)* |  |

*Cohen's Kappa = 0.20, $p < 0.001$*
*Note: VIA result was not reported for one participant, and HPV testing was not performed for one participant*

### Histology Findings

All 309 participants underwent colposcopy and biopsy. Histological examination revealed that the majority of participants (275, 89.0%) had no evidence of cervical intraepithelial neoplasia. Among those with abnormal histology, 9 (2.9%) had LSIL/CIN1, 15 (4.9%) had CIN2, 5 (1.6%) had CIN3, and 2 (0.7%) had cervical cancer. A total of 34 participants (11.0%) had histologically confirmed abnormalities, with 22 participants (7.1%) having CIN2+ lesions (CIN2, CIN3, or cancer) (Table 3).

**Table 3:** Histological Findings among All Participants (n=309)

| **Histological Finding** | **Frequency** | **Percentage** |
| --- | --- | --- |
| Normal | 275 | 89.0 |
| LSIL/CIN1 | 9 | 2.9 |
| CIN2 | 15 | 4.9 |
| CIN3 | 5 | 1.6 |
| Cervical cancer | 2 | 0.7 |
| **CIN2+ (CIN2, CIN3, Cancer)** | **22** | **7.1** |

### Comparison of HPV and VIA by Histology

Among participants with histologically confirmed CIN2+ (n=22), 20 were HPV-positive (90.9%) and 2 were HPV-negative (9.1%). In contrast, VIA was positive in only 10 of the 22 CIN2+ cases (45.5%), missing 12 cases (54.5%). Among the 287 participants without CIN2+, HPV was positive in 71 (24.7%), while VIA was positive in 26 (9.1%) (Table 4).

**Table 4:**
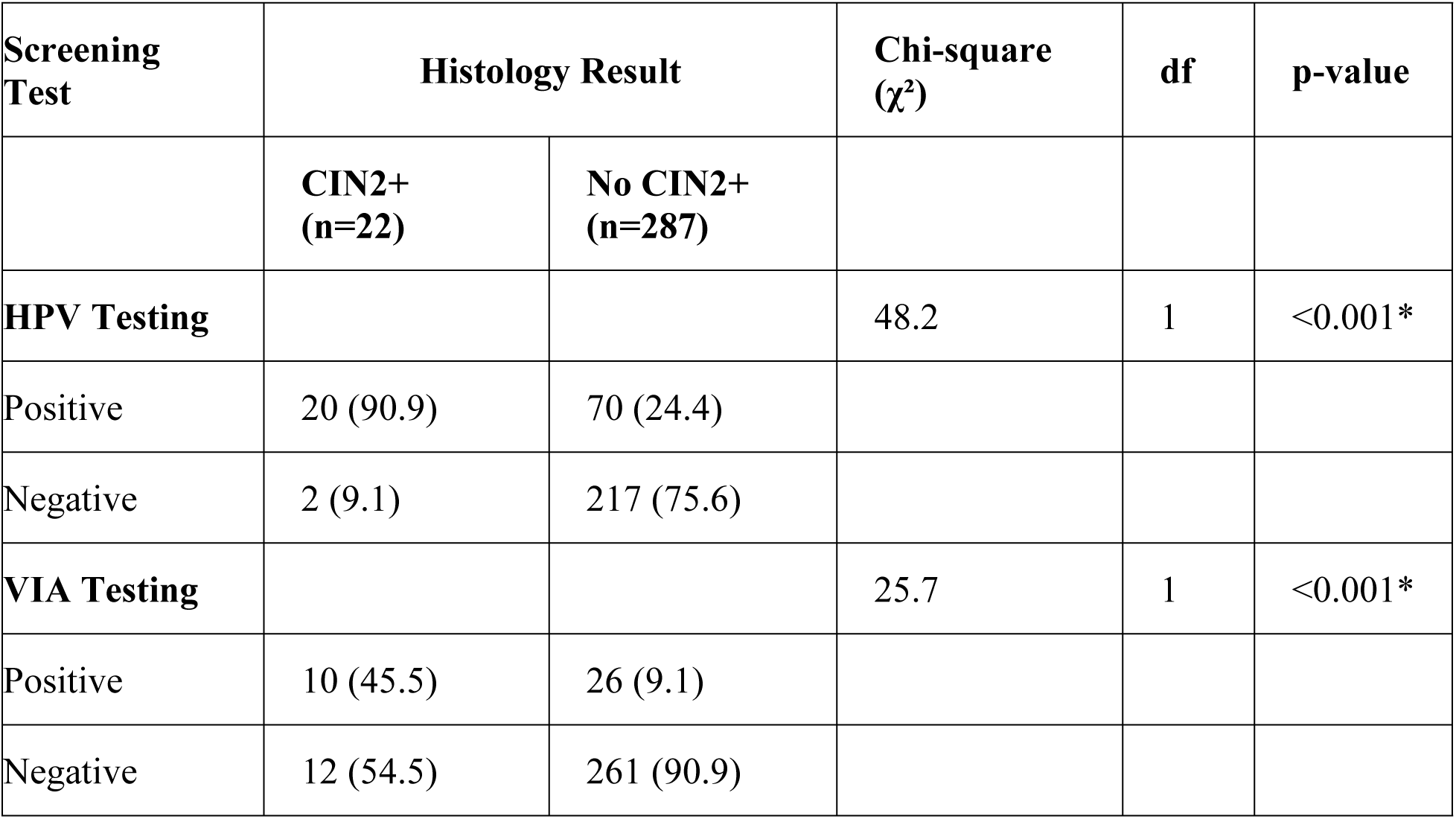
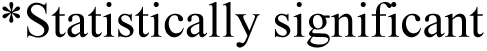
Distribution of Histology Results by HPV and VIA Status.

| **Screening Test** | **Histology Result** |  | **Chi-square ($\chi^2$)** | **df** | **p-value** |
| --- | --- | --- | --- | --- | --- |
|  | **CIN2+ (n=22)** | **No CIN2+ (n=287)** |  |  |  |
| **HPV Testing** |  |  | 48.2 | 1 | <0.001* |
| Positive | 20 (90.9) | 70 (24.4) |  |  |  |
| Negative | 2 (9.1) | 217 (75.6) |  |  |  |
| **VIA Testing** |  |  | 25.7 | 1 | <0.001* |
| Positive | 10 (45.5) | 26 (9.1) |  |  |  |
| Negative | 12 (54.5) | 261 (90.9) |  |  |  |

\*Statistically significant

The distribution of histology results by combined HPV and VIA status showed that HPV-positive/VIA-negative participants had the highest proportion of CIN2+ (12/69, 17.4%), followed by HPV-positive/VIA-positive participants (5/21, 23.8%). Notably, 2 cases of CIN2+ occurred in HPV-negative/VIA-positive participants, and no CIN2+ was detected in HPV-negative/VIA-negative participants (Table 4).

### Diagnostic Accuracy

With complete histological verification of all 309 participants, HPV testing demonstrated significantly higher sensitivity than VIA for detecting CIN2+ (91.2% [95% CI: 76.3-98.1] vs. 44.1% [95% CI: 26.7-62.6]; p<0.001). However, VIA showed higher specificity than HPV testing (91.7% [95% CI: 87.8-94.7] vs. 83.3% [95% CI: 78.4-87.6]; p=0.003). The positive predictive value was 37.4% (95% CI: 27.5-48.1) for HPV testing compared to 41.7% (95% CI: 25.5-59.2) for VIA (p=0.36). The negative predictive value was significantly higher for HPV testing than for VIA (98.9% [95% CI: 96.3-99.8] vs. 92.0% [95% CI: 88.0-95.0]; p<0.001). The positive likelihood ratio was similar for both tests (HPV: 5.5 [95% CI: 4.1-7.3]; VIA: 5.3 [95% CI: 3.0-9.4]), while the negative likelihood ratio was significantly better for HPV (0.11 [95% CI: 0.04-0.29] vs. 0.61 [95% CI: 0.42-0.87]) (Table 5).

**Table 5:** Diagnostic Performance of HPV and VIA Testing for CIN2+ (n=309)

| **Diagnostic Measure** | **HPV Testing (95% CI)** | **VIA Testing (95% CI)** | **Test Statistic** | **p-value** |
| --- | --- | --- | --- | --- |
| Sensitivity | 91.2 (76.3-98.1) | 44.1 (26.7-62.6) | McNemar's $\chi^2 = 12.5$ | **<0.001** |
| Specificity | 83.3 (78.4-87.6) | 91.7 (87.8-94.7) | McNemar's $\chi^2 = 9.1$ | **0.003** |
| Positive Predictive Value | 37.4 (27.5-48.1) | 41.7 (25.5-59.2) | $\chi^2 = 0.52$ | 0.36 |
| Negative Predictive Value | 98.9 (96.3-99.8) | 92.0 (88.0-95.0) | $\chi^2 = 14.8$ | **<0.001** |
| Positive Likelihood Ratio | 5.5 (4.1-7.3) | 5.3 (3.0-9.4) | — | 0.87 |
| Negative Likelihood Ratio | 0.11 (0.04-0.29) | 0.61 (0.42-0.87) | — | **<0.001** |
| Area under ROC Curve | 0.92 (0.87-0.96) | 0.70 (0.61-0.79) | $z = 4.8$ | **<0.001** |

The area under the receiver operating characteristic (ROC) curve was 0.92 (95% CI: 0.87-0.96) for HPV testing and 0.70 (95% CI: 0.61-0.79) for VIA, indicating superior overall diagnostic performance for HPV testing (p<0.001).

## Discussion

This study evaluated the diagnostic accuracy of VIA compared to HPV DNA testing for cervical cancer screening among women living with HIV in Osogbo, Nigeria, with the important methodological strength of complete histological verification of all participants. We found an HPV prevalence rate of 29.4% and a VIA-positivity rate of 11.7% in this population. The HPV prevalence is comparable to the 23.6% reported in Ibadan [22] but lower than the 36.5% reported in Lagos [23] and lower than the 33.3% reported in Brazil among WLHIV [21]. However, it is higher than the 9.1% reported in a community in China [24]. This variation may be attributed to differences in population characteristics, as most participants in our study were aged 40 years or older, married, and multiparous. Age has been shown to have a negative association with HPV infection rates, with older women demonstrating lower HPV prevalence irrespective of immune status [25]. The higher mean age of our participants (42.6 years) compared to other studies may partially explain the lower HPV prevalence observed.

The immunosuppression caused by HIV infection considerably influences the prevalence of HPV infection, with up to a six-fold increased risk of CIN [20]. The relatively low HPV prevalence in our study among WLHIV is likely due to the preponderance of older women and the fact that 92.6% of participants were on highly active antiretroviral therapy (HAART) compared to a study from a similar setting [26]. Earlier studies have reported increased clearance rates and reduction in high-risk HPV infection persistence in women living with HIV on treatment [27]. This may be attributed to the low HIV viral load observed in most participants receiving HAART. This concurs with a meta-analysis that reported an adjusted odds ratio of 0.83 (95% CI: 0.70-0.99) for high-grade CIN among WLHIV on ART, after adjusting for CD4 cell count and ART duration [28].

The HPV prevalence rate was significantly higher than the CIN prevalence confirmed by histology in this population. Approximately 7.1% (22/309) of our study participants had high-grade cervical intraepithelial neoplasia (CIN 2 and 3) confirmed by histology. This is lower than the 14.3% reported by Ezechi et al. [29] among WLHIV in Southwestern Nigeria. The low prevalence of confirmed high-grade CIN in our study may be attributed to the high proportion of participants on HAART, which has been shown to reduce the progression of CIN to higher grades.

Comparison of HPV testing with VIA showed that approximately one-quarter of those who were VIA-negative were HPV-positive (25.3%), while 15 of 218 HPV-negative participants (6.9%) had positive VIA results. The latter observation is noteworthy and warrants further discussion. Possible explanations for VIA positivity in HPV-negative women include the detection of other cervical pathologies such as cervicitis, cervical scarring, fibrosis, or the presence of low-risk HPV types not detected by the test. Additionally, VIA may detect acetowhite changes resulting from inflammatory conditions or metaplastic epithelium, which can lead to false-positive results. This highlights a limitation of VIA in terms of specificity when used as a standalone screening test. However, the complete histological verification in our study allowed us to determine that these VIA-positive/HPV-negative cases did not represent true CIN2+, confirming the lower sensitivity of VIA.

The congruence between the two tests was statistically significant (p=0.001), but the level of agreement was only fair, with a Cohen’s Kappa of 0.20. This suggests that while VIA and HPV testing are both useful screening tools, they identify different subsets of women at risk. This moderate-to-fair agreement is consistent with earlier studies that have reported decreased sensitivity of VIA in diagnosing CIN [10, 30]. Singh et al. similarly reported a low agreement level between HPV and VIA, which may be due to differences in the study population. Our findings suggest that VIA may be more reliable for screening WLHIV on HAART than in the general population, given the lower prevalence of HPV-related lesions in this treated population.

The diagnostic performance analysis, with the important strength of complete histological verification, revealed that HPV testing had significantly higher sensitivity (91.2%) compared to VIA (44.1%) for detecting CIN2+. This finding is consistent with the understanding that HPV testing is a more sensitive screening tool for identifying women at risk of cervical precancerous lesions. The high sensitivity of HPV testing means that it will miss far fewer cases of CIN2+ compared to VIA. In our study, VIA would have missed 54.5% of CIN2+ cases (12 of 22), while HPV testing would have missed only 9.1% (2 of 22). This has important clinical implications, as missed high-grade lesions represent opportunities for early intervention and prevention of cervical cancer.

The specificity of HPV testing was 83.3% compared to 91.7% for VIA, indicating that VIA produced fewer false-positive results than HPV testing. However, the clinical significance of this difference must be considered in the context of the screening program. In a resource-limited setting, false-positive results lead to unnecessary colposcopy referrals and patient anxiety, but false-negative results (missed cases of CIN2+) carry the risk of progression to invasive cervical cancer. Given the high mortality associated with cervical cancer in Nigeria, the superior sensitivity of HPV testing likely outweighs the benefit of higher specificity with VIA.

The positive predictive value was similar for both tests (37.4% for HPV vs. 41.7% for VIA), meaning that approximately 37-42% of women with positive screening tests would have confirmed CIN2+ on histology. This relatively low PPV reflects the high prevalence of transient HPV infections that do not progress to cervical lesions, particularly in this WLHIV population. The negative predictive value was significantly higher for HPV testing (98.9%) compared to VIA (92.0%), meaning that a negative HPV test virtually rules out CIN2+, while a negative VIA test still leaves an 8% chance of having CIN2+. This is a critical advantage of HPV testing, as a negative result provides substantial reassurance and reduces the need for repeat screening in the short term.

The likelihood ratios further support the superior performance of HPV testing. While both tests had similar positive likelihood ratios (5.5 for HPV vs. 5.3 for VIA), indicating that a positive result moderately increases the probability of CIN2+, the negative likelihood ratio for HPV (0.11) was much better than for VIA (0.61). A negative likelihood ratio of 0.11 means that a negative HPV test substantially reduces the probability of CIN2+, while a negative VIA test has a more modest effect. The area under the ROC curve confirmed that HPV testing has significantly better overall diagnostic accuracy (0.92 vs. 0.70 for VIA).

### Comparison with Previous Studies

The findings of this study are consistent with the growing body of evidence supporting HPV testing as the preferred primary screening method for cervical cancer. A systematic review and meta-analysis by Kelly et al. [9] found that HPV testing had higher sensitivity than VIA for detecting CIN2+ among WLHIV, with a summary sensitivity of 90% for HPV compared to 65% for VIA. Our results align closely with these estimates. Similarly, studies from other African countries, including Tanzania [15] and Mozambique [20], have reported superior sensitivity of HPV testing compared to cytology or VIA.

However, our finding of relatively high specificity for VIA (91.7%) is somewhat higher than reported in some studies [10, 30]. This may be explained by the high proportion of women on HAART in our study, which may reduce the prevalence of inflammatory conditions that can cause false-positive VIA results. Additionally, the use of trained research nurses and quality assurance measures may have improved VIA specificity.

The importance of complete histological verification in our study cannot be overstated. Previous studies that only verified positive screening results (verification bias) have been shown to overestimate sensitivity and underestimate specificity [31]. Our design, with all 309 participants undergoing colposcopy and biopsy, provides unbiased estimates that more accurately reflect the true performance of these screening tests in clinical practice.

### Clinical Implications

These findings have important implications for cervical cancer screening programs in Nigeria and other LMICs. The superior sensitivity and negative predictive value of HPV testing support its use as the primary screening tool for WLHIV, as recommended by WHO [8]. However, the cost and infrastructure requirements of HPV testing remain significant barriers in resource-limited settings.

A sequential screening approach, where HPV testing is used as the primary screen followed by VIA triage for HPV-positive women, could offer an optimal balance between sensitivity and specificity. In this strategy, HPV testing would identify women at risk (high sensitivity), while VIA would help prioritize those with visible lesions for immediate treatment (improved PPV). HPV-positive/VIA-negative women could be followed up with repeat testing in 12 months, while HPV-positive/VIA-positive women could be offered immediate treatment with thermo-coagulation or cryotherapy. This approach would reduce the number of unnecessary colposcopy referrals while maintaining high sensitivity for detecting CIN2+.

For settings where HPV testing is not available or affordable, VIA remains a viable alternative. The specificity of VIA in our study was good (91.7%), and it remains an inexpensive, low-technology option that can be performed by nurses and other healthcare workers. The “screen-and-treat” approach with VIA has been shown to be effective in reducing cervical cancer incidence and mortality in LMICs [11]. However, the lower sensitivity of VIA (44.1%) means that many women with CIN2+ will be missed, and programs using VIA alone should have robust follow-up mechanisms to ensure women with negative VIA results are re-screened at appropriate intervals.

### Strengths and Limitations

This study has several notable strengths. First, and most importantly, we achieved complete histological verification of all 309 participants, eliminating verification bias and providing unbiased estimates of diagnostic accuracy. Second, the blinding of colposcopists and pathologists to screening results minimized interpretation bias. Third, the study population was representative of WLHIV receiving care in Nigerian hospitals, enhancing generalizability. Fourth, the use of standardized training and quality assurance measures for VIA improved the reliability of this operator-dependent test.

### Limitations

HIV viral load or CD4 count was not assessed, which could influence HPV prevalence and CIN progression. These factors may have confounded the relationship between screening test results and histological outcomes. The cross-sectional design does not allow assessment of the long-term outcomes of screening, such as the prevention of invasive cervical cancer or mortality reduction.

## Conclusion

HPV DNA testing demonstrated significantly higher sensitivity and negative predictive value compared to VIA, while VIA showed higher specificity. The superior sensitivity of HPV testing means that it will miss far fewer cases of CIN2+, making it the preferred primary screening tool for WLHIV. However, VIA remains a valuable alternative in settings where HPV testing is not available, particularly given its higher specificity and lower cost. A sequential screening approach combining HPV testing as the primary screen followed by VIA triage may offer the optimal balance between sensitivity and specificity in resource-limited settings, reducing unnecessary colposcopy referrals while maintaining high sensitivity for detecting CIN2+.

Given the high burden of cervical cancer among WLHIV in Nigeria and other LMICs, urgent action is needed to implement effective screening programs. The WHO global strategy for cervical cancer elimination provides a framework for action, and our findings support the use of HPV testing as the primary screening method to achieve the 70% screening target by 2030. Future research should evaluate the cost-effectiveness of different screening strategies, assess the impact of HAART on screening performance, and develop implementation strategies tailored to resource-limited settings.

## Data Availability

The datasets generated and/or analyzed during the current study are not publicly available due to the sensitive nature of the data collected from women living with HIV and restrictions imposed by the Research Ethics Committee of Osun State University Teaching Hospital to protect participant privacy and confidentiality.

## Acknowledgments

We thank the staff of the Gynaecologic Oncology Research Laboratory of Obafemi Awolowo University Teaching Hospitals Complex, Ile-Ife, for their technical support. We also thank the management and staff of UNIOSUN Teaching Hospital and State Specialist Hospital, Asubiaro, Osogbo, for providing the facilities and resources necessary for this study. We are grateful to the research nurses and colposcopists who performed the screening examinations and colposcopies. Above all, we sincerely thank all the women who participated in this study for their time, trust, and cooperation, without whom this research would not have been possible.

## Ethics approval and consent to participate

This study was conducted in accordance with the Declaration of Helsinki. Ethical approval was obtained from the Research Ethics Committee of UNIOSUN Teaching Hospital, Osogbo, Nigeria (Approval Number: UTH/REC/2022/05/17/603). Written informed consent was obtained from all participants prior to enrollment in the study. Participants were informed of their right to withdraw from the study at any time without prejudice to their medical care. All data were anonymized and stored securely to ensure participant confidentiality.

## Consent for publication

Not applicable

## Author’s contribution

Bola-Oyebamiji and Awowole conceived and designed the study. Bola-Oyebamiji, Oyeniran, Bamkefa and Adekanle contributed to data collection. Adeyemo and Yussuf provided colposcopy and histology oversight. Oyeniran, Olabode, and Bola-Oyebamiji performed data analysis and interpretation. Bola-Oyebamiji Adeyemo and Olabode drafted the manuscript. All authors critically reviewed, revised, and approved the final version of the manuscript and agree to be accountable for all aspects of this work.

